# A Comparative Analysis of Signal Quality and Morphological Fidelity in ECG Smartwatches

**DOI:** 10.64898/2026.09.09.26362599

**Authors:** Safwan Mohammed, Sameh Sowelam, Nada Omran

## Abstract

This study provides a direct objective comparison of the ECG signal quality and morphological features preservation of four leading commercial smartwatches: the Apple Watch Series 9, Samsung Galaxy Watch 6, Fitbit Sense 2, and Withings ScanWatch, using a controlled dataset. The Philips TC30 is used as the reference standard. ECG signal quality is assessed through Signal-to-Noise Ratio (SNR), Standard Deviation of Baseline Wander and Power Spectral Density (PSD), while the preservation of the morphological features is assessed through comparing the performance differences of our J-point and ST deviation automatic detection algorithms when run on each watch’s recordings separately. J-point detection is assessed by comparing each device’s performance against the reference device, while ST deviation is assessed using simulator defined ground truth values. The Apple Watch and Withings ScanWatch had the best SNR, while the Samsung Galaxy Watch had much lower values. The Withings ScanWatch had the most stable baseline, while the Fitbit Sense 2 had the most variable baseline. All devices showed a consistent negative bias in J-point detection, which suggests that internal signal processing tends to smooth out small changes in waveforms. For ST-segment classification, the Apple Watch achieved the most balanced performance across sensitivity and specificity. These results give researchers and clinicians data-driven advice on how to choose the best device for their specific needs.

## I. Introduction

On designing and conducting clinical trials, the gold standard for collecting physiological data has traditionally been the in-lab assessment. However, these measurements provide only a limited time window, potentially missing critical events and failing to capture the natural variability of human physiology in daily life [1] [2]. Wearable technology has the potential to overcome this challenge by enabling continuous and unobtrusive monitoring [2]. Since cardiovascular diseases remain a leading cause of morbidity and mortality globally [3], cardiac health biomarkers are among the highest priority parameters that require precise monitoring and analysis. Many cardiac conditions are serious, sudden, and require early detection and intervention [3]. This clinical need has resulted in a competitive market, encouraging numerous companies to develop and enhance smart wearable devices capable of recording electrocardiogram (ECG) snippets. As a result, a wide variety of devices is now available, each offering different combinations of capabilities and performance characteristics [4]. Despite the benefits of this variety for innovation, it also creates a significant challenge in determining which device best suits a particular study or clinical requirement. When choosing the optimal wearable to use, one must choose between many factors based on the study requirements such as: cost, battery life, device placement, signal quality, measurement accuracy, weight, size, and usability [5]. Although the relative importance of these factors varies across studies, signal quality and measurement accuracy are particularly critical in cardiac health monitoring. A fundamental challenge of using wearables in an uncontrolled environment is the high susceptibility to signal noise and motion artifacts, especially for signals requiring stationary measurement, like the ECG [6]. To accurately assess the wearables ECG signal quality and their ability to capture features of interest, one must have a clear understanding of the ECG characteristics. The ECG is a time varying bioelectrical signal with well-defined morphological features that reflect the electrical activity of the heart. Its characteristic waveform consists of distinct components, including the P wave, QRS complex, and T wave, as well as clinically important segments and points such as the ST segment and J-point [6]. Each of these features occupies a specific temporal and frequency range, with the majority of diagnostically relevant ECG energy concentrated below 40 Hz. Very low frequency components such as baseline wander are generally considered artifacts rather than features of clinical interest. High frequency noise may arise from muscle activity (electromyographic interference), powerline interference, electronic circuitry, and motion-induced electrode impedance changes [6]. These noise sources can distort wave-form morphology and critical features, therefore reduce the reliability of automated detection algorithms. Consequently, analyzing the ECG in both the time and frequency domains is essential for understanding how well a wearable device preserves clinically meaningful information. This paper addresses the challenge of device selection by providing a direct, objective comparison of the signal quality of leading commercial smartwatches: the Apple Watch Series 9, Samsung Galaxy Watch 6, Fitbit Sense 2, Withings ScanWatch, using a controlled dataset [7]. The paper also addresses the usability of these commercial wearable devices for diagnostic purposes derived from the morphological features of the recorded ECG signal. The clinical-grade Philips TC30 device is used as the reference standard for agreement-based performance evaluation. The paper begins with an assessment of fundamental signal quality metrics, including Power Spectral Density (PSD), baseline wander, and Signal-to-Noise Ratio (SNR). We then evaluate the impact of signal quality on clinically relevant feature extraction, focusing on J-points and ST-segment elevation and depression events. For each feature, a single automated detection algorithm is applied offline and uniformly to all recordings from all devices. J-point detection performance is primarily assessed in terms of agreement with the reference device. For ST-segment elevation and depression events, performance is evaluated relative to simulator-defined ground truth values. The findings of this analysis offer data-driven evidence to aid researchers and clinicians in selecting the most suitable wearable device for specific cardiovascular monitoring applications

## II. Methods

This study’s comparative analysis of wearable ECG devices was organized into three main parts: evaluating signal quality metrics (SNR and Baseline Wander), conducting spectral analysis (PSD), and assessing how well each device detects key ECG points (J-point and ST-segment errors compared to a reference standard).

### A. ECG Signal Acquisition and Dataset

We used dataset introduced at [7], it consists of 173 paired ECG recordings obtained from four commercial smartwatches: the Apple Watch Series 9, Samsung Galaxy Watch 6, Fitbit Sense 2, and Withings ScanWatch, as well as a hospital-grade ECG (Philips TC30) that was used as a reference device [7]. The Philips TC30 recordings were collected using standard calibrated ECG leads. Recordings were collected under several configurations, using a METRON PS-440 patient simulator, that include: variations in heart rate (30–300 beats per minute), R-wave amplitude (500 µV to 2000 µV), and ST-segment elevation and depression (−800 µV to 800 µV) [7]. This subset of recordings provided a balanced dataset suitable for robust signal quality characterization and statistically meaningful comparisons using both categorical (McNemar’s test) and paired nonparametric analyses.

### B. Signal Quality Assessment

Two commonly used metrics were used to measure each device’s signal quality and noise characteristics: SNR and the standard deviation of Baseline Wander.

#### 1) Signal-to-Noise Ratio (SNR)

Each recording was first segmented into individual cardiac cycles by dividing the trace between successive R-peaks. R-peaks were detected using the Pan-Tompkins algorithm [8] Fig.1. These cycles were then zero padded to the length of the maximum cycle to account for heart rate variability and ensure uniform dimensionality for vector operations. Then for each segment, its cosine similarity to every other segment was computed after determining and applying the optimal temporal shift that maximizes this similarity. Determining the optimal shift was done through iteratively shifting the segment within a bounded window and the shift achieving maximum cosine similarity was retained. The segment with the highest mean of these maximized similarities was selected as the master template. All remaining segments were then aligned to the selected template using the same shift-search algorithm, maximizing cosine similarity defined as

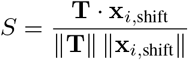

where **T** denotes the template and **x**_*i*,shift_ the temporally shifted segment. Segments achieving a similarity *S* ≥ 0.8 were retained. A single averaged segment was computed from all the retained aligned segments. The signal power (*P*_signal_) was then defined as the mean square of this averaged segment. The noise component was estimated by subtracting this averaged segment from each individual aligned cardiac cycle, and the noise power (*P*_noise_) was calculated as the mean square of the noise component. The final SNR in decibels (dB) was computed as

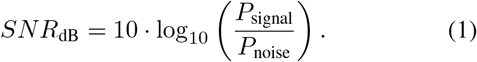

**Fig. 1:**
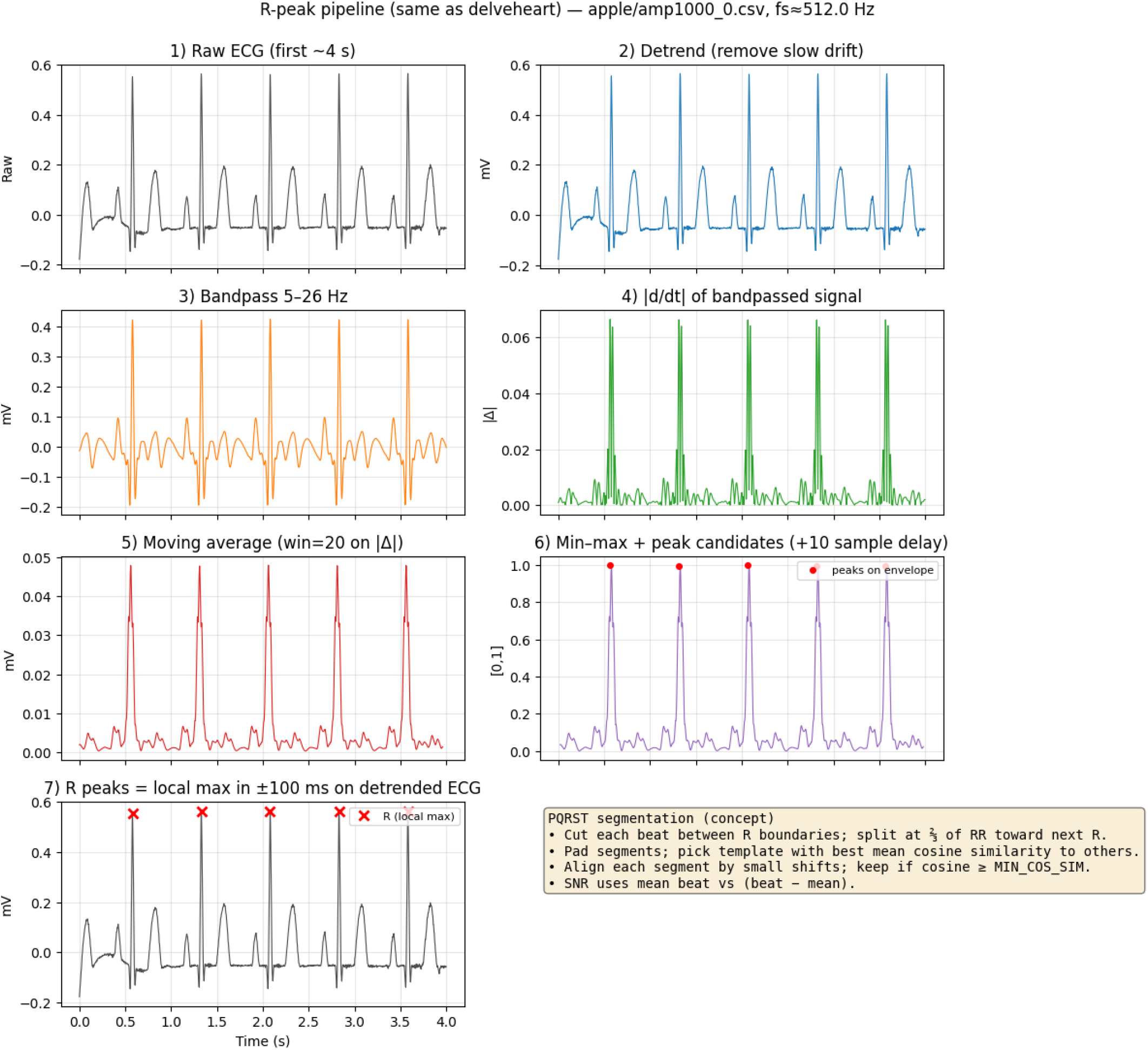
Example of template alignment process.

#### 2) Baseline Wander Standard Deviation (BW Std)

Base-line wander (BW) was estimated using the two-stage cascading median filtering approach described by [9] [10]. This method is specifically designed to isolate the low-frequency baseline by sequentially suppressing the various morphological components of the ECG signal. The raw ECG signal was first processed with a moving median filter with a window size of 200 ms. This first stage effectively removes the high-frequency QRS complexes and P waves. The resulting signal was then subjected to a second median filter with a window size of 600 ms, which suppresses the remaining T waves. The output of this second filtering stage represents the isolated baseline of the ECG signal [9]. The standard deviation (Std) of the extracted baseline signal was computed for each recording. This value quantifies the average amplitude of the baseline fluctuation across the entire recording.

### C. Spectral Analysis

#### Power Spectral Density (PSD)

The Power Spectral Density (PSD) characterizes how the power of the ECG signal and noise are distributed across different frequencies. This analysis is vital for identifying common noise sources (e.g., 50*/*60 Hz power-line interference, low-frequency BW) and for understanding the effective bandwidth of each device. The

PSD was computed using Welch’s method [11] with a Hann window. A segment length of approximately 8 seconds (or the maximum possible length for shorter recordings) with a 50% overlap was employed to balance frequency resolution and variance reduction. To enable a comparative device-level analysis, a common frequency grid was established for each smartwatch based on its nominal sampling frequency. Individual PSD estimates were interpolated onto this uniform grid. Finally, for each device, for each data sample, we subtracted the PSD of the reference from the wearable’s PSD to obtain the deviation of the PSD from the reference, then for each device, mean and std across samples was observed, see Fig.4.

### D. Signal Quality Effect on Fiducial Point Detection Performance

To evaluate how differences in ECG signal quality across wearable devices affect automated fiducial point extraction, the performance of J-point detection and ST-segment morphology estimation was analyzed. A unified automated detection algorithm was applied to all recordings, ensuring that observed performance differences were attributable to signal quality variations rather than algorithmic inconsistencies.

#### 1) Fiducial Point Extraction Procedure

Each ECG recording was segmented into individual cardiac cycles using R-peak detection. Beats deemed physiologically valid based on duration and morphology constraints were retained. The retained segments were then temporally aligned using the same ensemble-alignment procedure described in Section II-B1, including zero-padding to a common length and template-based shift alignment. Following alignment, a beat template was constructed by averaging all aligned cardiac cycles, thereby reducing random noise while preserving consistent fiducial morphology.

Following template construction, the P wave, S wave, and T wave were segmented using a single-atom Gaussian matching approach, inspired by matching pursuit methods [12].

For each waveform (P, S, and T), an independent search was performed within physiologically constrained time windows relative to the R-peak [13]. Within each window, a single Gaussian atom was fitted by maximizing the normalized correlation between the detrended signal segment and the candidate atom.

In cases where no atom met the predefined correlation and amplitude criteria, the corresponding J-point or ST-segment output was marked as missing. This was followed by manual validation by experts using internally defined UI tools to ease the process.

#### 2) J-Point Detection Agreement Analysis

The J-point was defined as the terminal point of the segmented S wave obtained from the Gaussian atom decomposition. This definition aligns with the physiological interpretation of the J-point as the transition between ventricular depolarization and repolarization [14].

The effect of signal quality on J-point detection behavior was evaluated using paired agreement analysis relative to the reference device. J-point detections from a smartwatch and the reference device were considered concordant if both devices detected a J-point and the absolute timing difference satisfied:

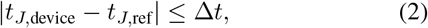

where Δ*t* = 30 ms accounts for expected temporal variability due to wearable sensor placement, bandwidth limitations, and residual noise.

Detection outcomes were categorized per recording as detected or not detected, yielding paired binary outcomes for each device–reference comparison. For each device detection behavior was assessed using McNemar’s test [15], which evaluates asymmetric disagreement between paired binary classifiers. This test determines whether signal degradation introduces systematic detection differences relative to the reference ECG.

For recordings in which both devices detected a J-point within the tolerance window, the J-point timing error was analyzed using the Wilcoxon signed-rank test [16] to assess the presence of systematic temporal bias in J-point localization attributable to signal quality differences.

#### 3) ST-Segment Deviation and Morphology Analysis

ST-segment deviation was quantified on the averaged beat template as the difference between the mean T-wave baseline and the mean P-wave baseline. For each recording, the ST deviation value was computed as:

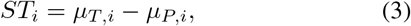

where *µ*_*T,i*_ and *µ*_*P,i*_ denote the estimated baseline levels of the T and P waves, respectively.

To classify ST morphology, a constant amplitude-based threshold was applied uniformly across all devices. The threshold was defined as:

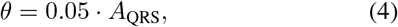

where *A*_QRS_ is the global amplitude of the QRS complex measured on the averaged template. An ST segment was classified as elevated if *ST*_*i*_ *> θ*, depressed if *ST*_*i*_ *<* − *θ*, and normal otherwise.

For each smartwatch, categorical ST detection performance was evaluated independently against simulator-defined ground truth values. Separate binary classification analyses were performed for ST elevation and ST depression. Confusion matrices were computed for each device, from which accuracy, sensitivity, specificity, and precision were derived. Recordings with missing or invalid ST estimates were treated as negative detections for the corresponding class.

## III. Results

### A. Signal Quality Assessment

#### 1) Signal-to-Noise Ratio (SNR)

The Apple Watch and Withings ScanWatch exhibited the highest mean SNR values (21.97 ± 3.31 dB and 21.85 ± 2.67 dB, respectively), followed by the Fitbit Sense 2 (18.18 ± 3.32 dB). The Samsung Galaxy Watch 6 demonstrated substantially lower SNR values, with a mean of 10.22 ± 2.45 dB.

The distribution of SNR values for each device is shown in Fig. 2a.

**Fig. 2:**
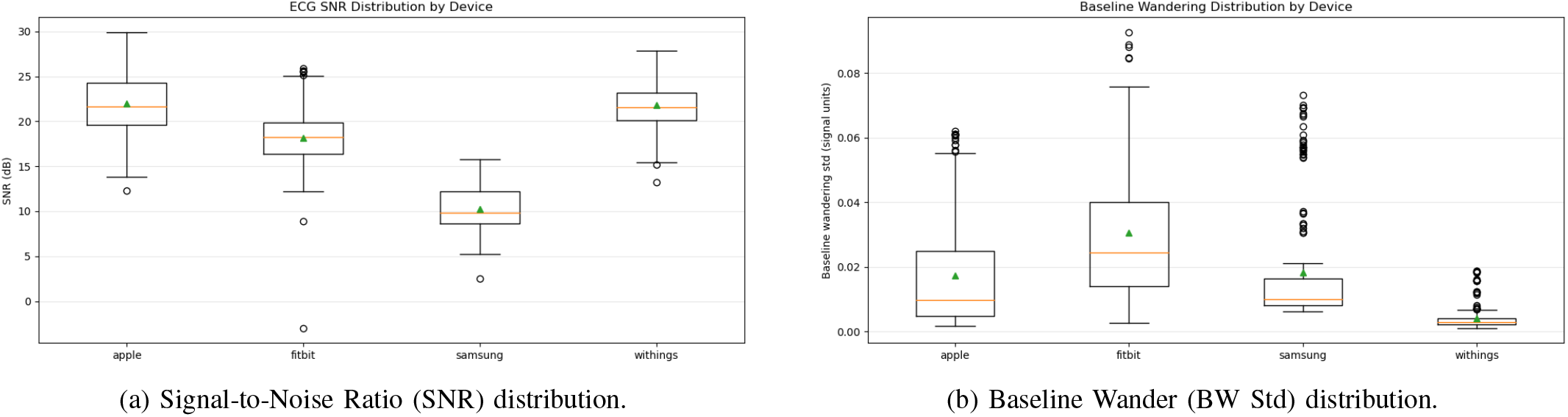
Distribution of signal quality metrics across devices. In the box plots, the orange horizontal line represents the median, while the green triangle indicates the mean.

#### 2) Baseline Wander Standard Deviation (BW Std)

The Withings ScanWatch exhibited the lowest baseline variability (mean 0.0042 ± 0.0038), while the Fitbit Sense 2 showed the highest baseline wander variability (mean 0.0307 ± 0.0215). Apple Watch and Samsung Galaxy Watch demonstrated comparable baseline wander levels (mean 0.0173 ± 0.018, mean 0.0184 ± 0.0184 respectively).

The distributions of baseline wander standard deviation across devices are shown in Fig. 2b.

### B. Spectral Analysis

By comparing the PSD response of the ECG sensor of each watch with that of the reference Philips device (Fig 3), Withings showed the least deviation from the reference, with minor zero-mean deviation at around 15 Hz, and slight negative deviation at very low frequency (0.5-1 Hz), which happened due to slow baseline wandering. Samsung showed the most deviation from the reference, with unbalanced deviation, that expanded on the spectrum (¡30 Hz). Apple showed a large magnitude, zero-mean deviation expanded from about (7-15 Hz). Fitbit had the same baseline wandering effect at low frequency (0.5-1 Hz), and a low-magnitude effect spread across (1 - 12 Hz).

**Fig. 3:**
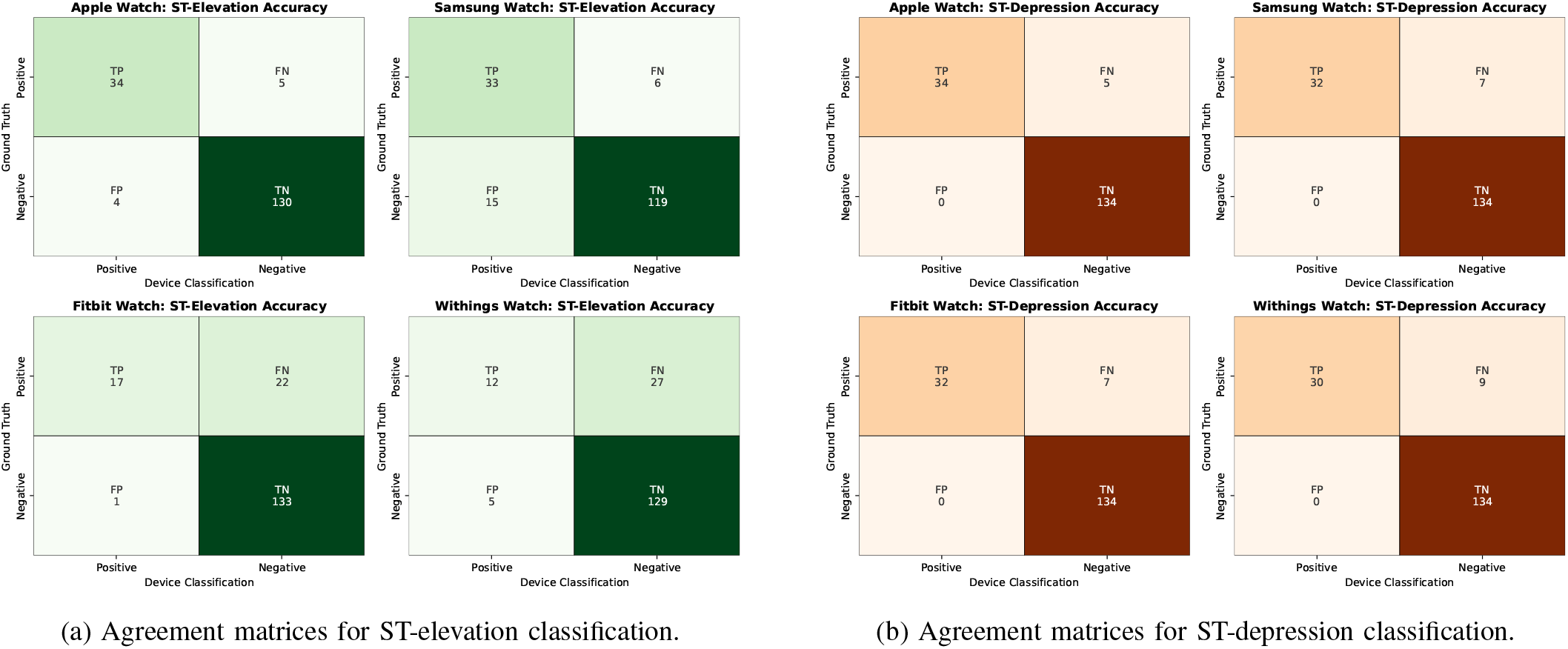
Paired agreement matrices for categorical ST-segment morphology classification across the four smartwatches.

**Fig. 4:**
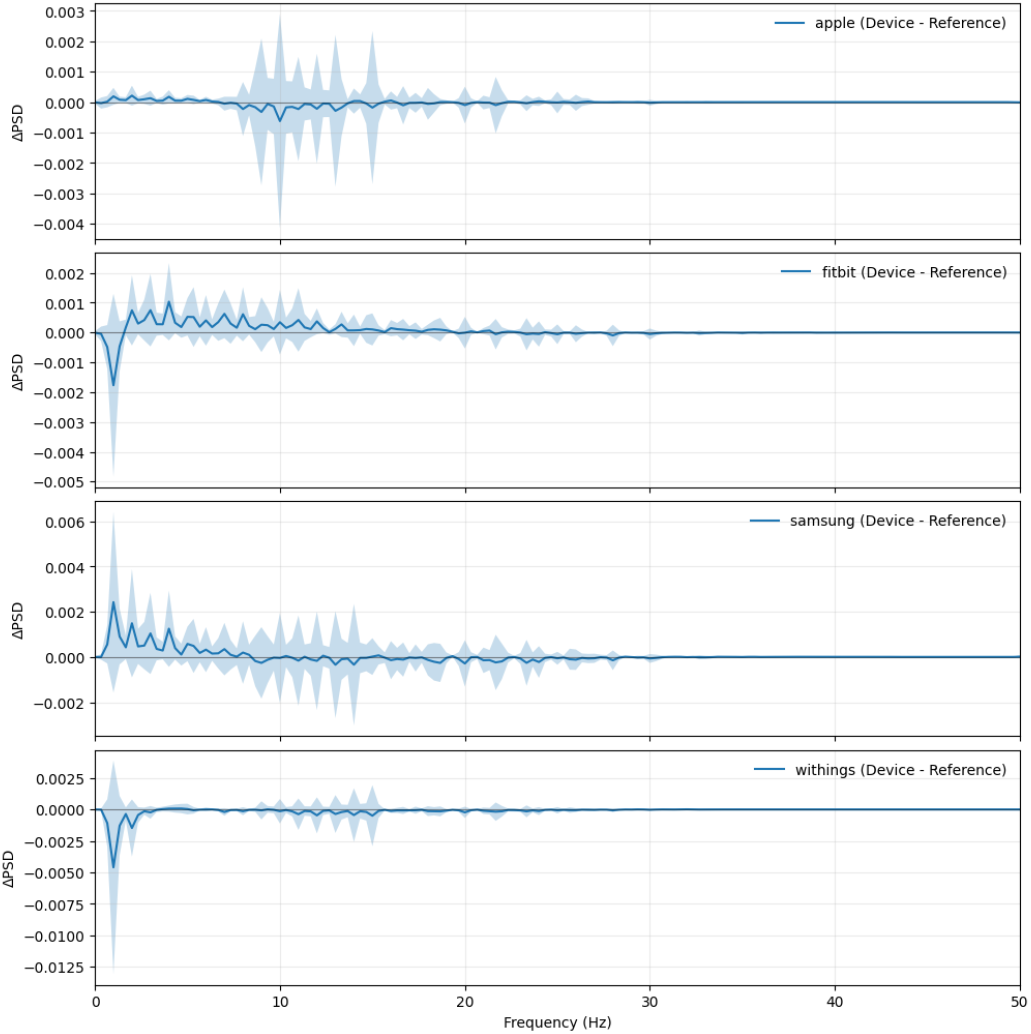
Deviation from the reference of the PSD for each device. Average: Solid blue line, Standard Deviation: soft shaded blue

### C. Signal Quality Effect on Fiducial Point Detection Performance

#### 1) J-Point Detection Performance

The comparative J-point detection performance of each device relative to the reference is summarized in Fig. 5. The Samsung Watch exhibited the lowest total number of disagreements with the reference, totaling 21 cases. Its McNemar test p-value (*p* = 0.07) indicates that those disagreements are likely due to chance rather than a significant systematic detection bias. All Smartwatches exhibited significant p-values (*p <<<* 0.001) and high negative biases (Apple: *bias* = −1.0, Samsung: *bias* = −0.619, Withings: *bias* = −1.0, Fitbit: *bias* = −0.9393).

**Fig. 5:**
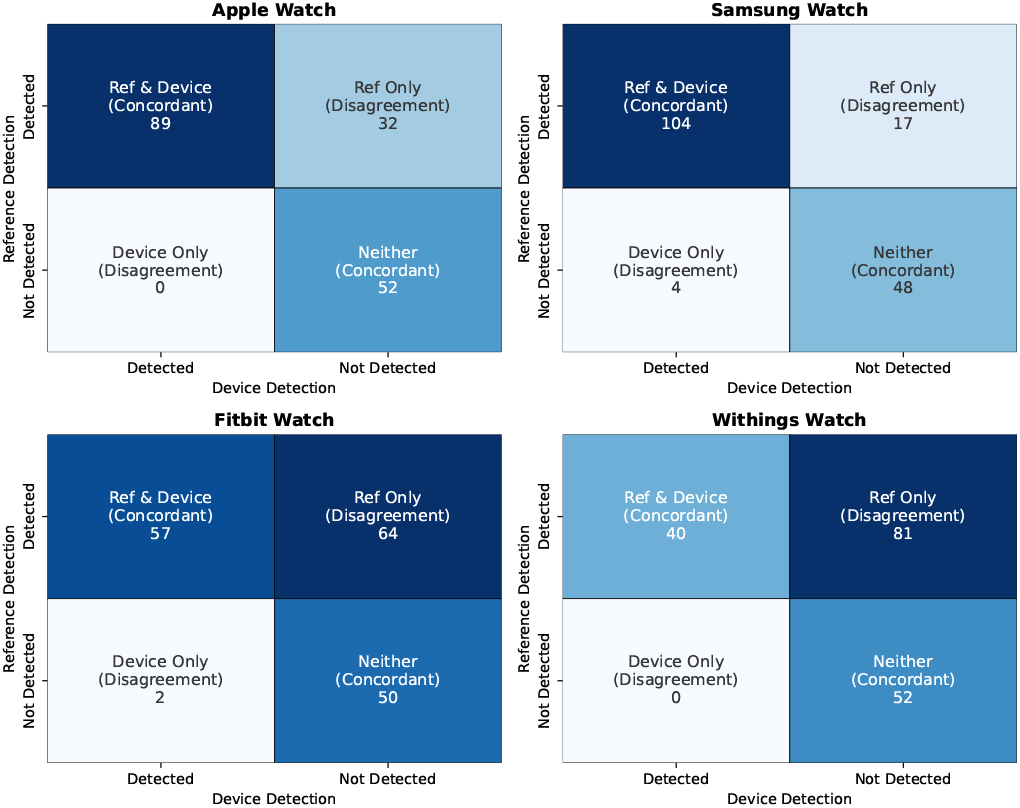
J-point detection agreement matrices for each smart-watch relative to the reference device.

For recordings where J-points were successfully detected by both the wearable and reference within the 30 ms tolerance, the temporal accuracy was evaluated. The median timing errors for the Apple Watch, Fitbit Sense 2, Samsung Galaxy Watch 6, and Withings ScanWatch were 6.8 ms, 13.999 ms, 4.0 ms, and 8.3 ms, respectively. In all cases, the Wilcoxon signed-rank test yielded *p <* 0.001, indicating a statistically significant, though clinically small, systematic temporal shift in J-point localization across all wearable devices.

#### 2) ST-Segment Morphology and Deviation Analysis

The ability of the devices to correctly classify ST-segment elevation and depression events is summarized in Fig. 3a and 3b. For each device, confusion matrices were computed and used to derive accuracy, sensitivity, specificity, and precision.

For **ST-segment elevation**, the Apple Watch achieved an accuracy of 94.79%, with a sensitivity of 87.17%, specificity of 97.01%, and precision of 89.47%. The Samsung Galaxy Watch showed an accuracy of 87.86% and a sensitivity of 84.61%, but lower specificity (88.8%) and precision (68.75%). The Fitbit achieved an accuracy of 86.7%, with high specificity (99.25%) and precision (94.44%), but markedly reduced sensitivity (43.58%). The Withings ScanWatch demonstrated an accuracy of 81.5%, sensitivity of 30.76%, specificity of 96.26%, and precision of 70.58%, reflecting limited elevation detection capability despite a low false positive rate.

For **ST-segment depression**, all devices achieved perfect specificity (100%) and precision (100%), indicating the absence of false positive depression detections. The Apple Watch Series 9 achieved the highest sensitivity (87.17%) and overall accuracy (97.1%). The Samsung Galaxy Watch 6 and Fitbit Sense 2 both demonstrated sensitivities of 82.05% and accuracies of 95.95%. The Withings ScanWatch exhibited a sensitivity of 76.92% and an accuracy of 94.79%. Across all devices, false negatives accounted for the primary source of classification error in depression detection.

## IV. Discussion

This study provides a comprehensive, data-driven comparison of signal quality and diagnostic feature preservation across four leading commercial smartwatches. By utilizing a controlled dataset following IEC 60601-2-25:2011 standards [17], we minimized external confounding variables, such as skin-to-electrode impedance changes or body movement, allowing for an isolated assessment of each device’s internal hardware and signal processing characteristics. Our results reveal a clear hierarchy in signal fidelity. The Apple Watch Series 9 and Withings ScanWatch emerged as leaders in Signal-to-Noise Ratio (SNR). High SNR is a critical prerequisite for clinical applications, as it ensures that the morphological features required for diagnosis are not obscured by circuit noise. Conversely, the Samsung Galaxy Watch 6 demonstrated a significantly lower SNR, highlighting the need for the integration of signal filtering and preprocessing techniques.

Baseline wander (BW) remains a challenge for wearable integration, typically stemming from the electrode-skin interface. The Withings ScanWatch showed superior stability in this regard, whereas the Fitbit Sense 2 exhibited the highest variability. High baseline wander is particularly problematic for ST-segment analysis, as slow-varying offsets can distort the apparent ST level and reduce sensitivity to subtle elevation or depression patterns. The implications of these signal quality differences were further explored through automated fiducial point and ST-segment analyses. J-point detection was evaluated through agreement-based comparisons with a hospital-grade ECG reference, given the absence of simulator-defined ground truth annotations for this feature. A key finding was the systematic negative bias in J-point detection observed in all devices. This indicates that even high-quality wearables tend to “over-filter” or smooth the signal. While this enhancement favors sharp, high-amplitude peaks like the R-wave, it may filter out subtle transitions in electrical activity like the J-point. The systematic temporal shifts identified in the Wilcoxon tests (4 to 13.9 ms) may be negligible during manual inspection. However, for automated ST-segment measurements, a 13.9 ms shift is critical, as many clinical algorithms assess ST deviation at J + 60 ms or J + 80 ms, so this shift will directly propagate error into the diagnostic measurement.

In contrast to J-point analysis, ST-segment elevation and depression detection was evaluated directly against simulator-defined ground truth, enabling absolute performance assessment that reflects the integrity of the underlying ECG signal. Because a single, fixed detection algorithm was applied uniformly across all devices, observed differences in classification performance are due to variations in signal quality and device-specific preprocessing rather than algorithmic effects.

For ST-segment elevation, the Apple Watch exhibited the most balanced performance, combining high sensitivity with high specificity. This suggests that its signal preserves both low-frequency ST morphology and sufficient amplitude resolution to enable reliable detection of moderate elevation levels. The Samsung Galaxy Watch demonstrated comparable sensitivity but reduced precision, indicating that noise or baseline instability may introduce false ST deviations that cross the detection threshold. In contrast, the Fitbit and Withings ScanWatch achieved high specificity but markedly reduced sensitivity, reflecting that elevation patterns are frequently attenuated or smoothed below the detection threshold. ST-segment depression detection exhibited uniformly high specificity across all devices, with no false positive detections observed. Performance differences were therefore driven primarily by sensitivity, which varied across devices. This pattern indicates that while baseline stability and low-frequency noise suppression were generally sufficient to prevent false depression artifacts, the ability to preserve small negative ST deviations was limited by signal attenuation and bandwidth constraints. Overall, the ST-segment analysis highlights the trade-offs inherent in wearable ECG acquisition. High specificity across devices suggests effective suppression of low-frequency artifacts, but reduced sensitivity particularly for subtle deviations.

Taken together, these results underscore the trade-offs inherent in wearable ECG design and provide practical guidance for selecting wearable devices based on the intended clinical or research context. In (Table 1), we provide a list of recommendations for use cases or each watch, based on the strength and weakness of each one.

**TABLE 1:** Summary of recommended use cases, limitations, and key weaknesses for each smartwatch.

| Wearable Device | Recommended Use Cases | Use Cases to Avoid / Limitations | Key Weakness |
| --- | --- | --- | --- |
| Apple Watch Series 9 | Ischemia & Repolarization abnormalities: Best for ST-segment elevation/depression detection due to high accuracy (94.79%+) and balanced sensitivity/specificity. | Highly precise timing: While strong, it still possesses a slight systematic negative bias in J-point localization. | Over-filtering: Tends to smooth subtle waveform transitions. |
| Withings ScanWatch | Long-term monitoring & Baseline stability: Ideal when the electrode-skin interface is unstable, as it had the most stable baseline. | Subtle ST Elevation: Very low sensitivity (30.76%) for detecting ST-segment elevation events. | Significant Attenuation: Fails to capture many elevation patterns, smoothing them below detection thresholds. |
| Samsung Galaxy Watch 6 | J-Point Analysis: Best for morphological markers like the J-point; it had the lowest total disagreements with the reference device. | Clinical Ischemia Detection: Low SNR and reduced precision in ST-segment analysis increase the risk of false positives. | Low Signal Quality: High levels of circuit noise or internal interference (lowest SNR at ~10 dB). |
| Fitbit Sense 2 | General Cardiac Rhythm: Suitable for basic monitoring where baseline shifts are not the primary metric. | ST-Segment Elevation: Marked reduction in sensitivity (43.58%) for elevation detection. | Baseline Wander: Highest variability in baseline stability, which can distort ST-level measurements. |

## V. Conclusion

This research demonstrates that while commercial smart-watches have made significant strides toward clinical-grade ECG monitoring, they are not interchangeable. Across the evaluated devices, the Apple Watch Series 9 consistently exhibited strong signal quality characteristics, including high SNR and balanced baseline stability, which translated into more reliable preservation of ST-segment morphology. The Withings ScanWatch demonstrated similarly strong performance in baseline stability and overall signal fidelity, though with reduced sensitivity to subtle ST deviations. The Samsung Galaxy Watch 6, despite lower SNR, maintained reasonable performance for J-point analyses. The Fitbit Sense 2 showed higher baseline variability, which was reflected in reduced sensitivity for certain ST-segment deviations.

In summary, researchers and clinicians must select a device based on their specific analytical needs. If the study relies on certain morphological markers like the J-point, a device with less aggressive signal smoothing may be preferable, even at the expense of a lower SNR. However, for clinical research focusing on ischemia or repolarization abnormalities, researchers should prioritize devices with higher SNR and proven stability in ST-segment classification. Future developments in wearable ECG should aim to balance noise suppression with the preservation of subtle but clinically vital waveform features.

## Data Availability

All data produced are available online at: https://physionet.org/content/ecg-capable-smartwatches/1.0.0/

## Acknowledgment

The authors acknowledge Delve Health for granting access to the Delve Toolbox, which was utilized for ECG data processing in this study.

